# Organizational Readiness for the Implementation of a Three-Month Short-Course TB Preventive Therapy Regimen (3HP) in Four Health Care Facilities in Zimbabwe in 2020: A Mixed Methods Study

**DOI:** 10.1101/2021.05.26.21256736

**Authors:** Dorothy T. Chisare, Rutendo B.L. Zinyama-Gutsire, Charles Chasela

**Affiliations:** Division of Epidemiology and Biostatistics, School of Public Health, Faculty of Health Sciences, University of the Witwatersrand, Johannesburg, South Africa; Immunology Department, College of Health Sciences, University of Zimbabwe, Harare, Zimbabwe; Implementation Science Unit Programme, Right to Care, Johannesburg, South Africa

**Keywords:** Organizational readiness, TB preventive therapy, TPT, 3HP, Zimbabwe

## Abstract

**Background:** Tuberculosis preventive therapy (TPT) for latent TB infection has had limited success in Zimbabwe. The country plans to roll-out the three-month short-course TPT regimen (3HP) to address the implementation lag and poor uptake of the 6-9-month regimens. The study measured the level of organizational readiness while identifying barriers and facilitators to implement 3HP in four health facilities in Zimbabwe.

**Methods:** A convergent, parallel mixed-methods approach was used to collect data from four primary healthcare clinics in Bulawayo and Harare Metropolitan provinces, Zimbabwe. Twenty healthcare providers completed a 35-item, self-administered questionnaire designed on a 5-point Likert scale and developed from the Weiner organizational readiness model. Nine of the providers and five TB program managers took part in 20-30 minute individual semi-structured key-informant interviews. Median scores with interquartile ranges were calculated wherein a score of 3.3 or greater indicated readiness. Differences between facilities were assessed using a Kruskal-Wallis rank test. Qualitative data on barriers and facilitators were transcribed and analyzed using a framework approach.

**Results:** Readiness to implement 3HP across the four facilities was positive with a score of 3.8(IQR 3.3-4.1). The difference between the best 4.0(IQR 3.8-4.2) and worst-performing facility 3.2(IQR 2.7-3.3) was 0.8 and statistically significant (p=0.039). The low facility score was due to poor contextual factors 2.5(IQR 2.0-3.3), task demands 2.6(IQR 2.3-2.9), and resource availability 2.1(IQR 1.5-2.5) scores. Key organizational readiness facilitators included healthcare provider and management buy-in; community willingness to generate demand for 3HP; strong collective capability through task-shifting, alignment with existing primary healthcare programs, perceived benefits, and need for 3HP. Barriers were negative past TPT experiences, suboptimal programmatic monitoring, inconsistent health provider remuneration, inadequate staffing, added workload, and an erratic supply chain across facilities. The organizational communication gap prompts the slow program implementation culture.

**Conclusions:** The varied scores between facilities suggest distinct underlying conditions for readiness. Healthcare provider motivation is temporary based on the inconsistent resource supply, absence of TPT-specific monitoring and evaluation, and daily contextual challenges in facilities that must be addressed. Similar research is necessary for countries yet to implement 3HP to optimize the design or revision of delivery strategies and increase uptake of TPT.

## Introduction

Despite significant increased access to treatment, tuberculosis (TB) accounted for 10 million active cases and 1.3 million deaths in 2018 globally [1]. The re-occurrence of the growing burden of active TB disease is driven by latent tuberculosis infection (LTBI), particularly in developing countries. Persons with LTBI harbour Mycobacterium tuberculosis bacilli but are not infectious and cannot spread TB to others [2], however, once infected, individuals are at the highest risk of developing active TB [2]. One-third of the global population are carriers of LTBI which is an estimated 2.3 billion possible new TB cases [2]. The WHO Africa, Western Pacific, and South-East Asia regions have the highest prevalence of LTBI and account for 80% of the cases [2]. Zimbabwe fosters a high LTBI rate of 210 per 100 000 population and is one of 12 countries that contribute to 50% of the global TB burden [3, 4]. With 14% of the population living with HIV, more individuals are at the risk of LTBI wherein progression to active TB is high [3, 4].

As per WHO, a crucial strategy to prevent and eliminate active TB cases has been the treatment of LTBI through the provision of TB preventive therapy (TPT), specifically daily self-administered isoniazid preventive therapy (IPT)[3]. Provision of IPT is marred with challenges including constrained effectiveness due to toxicity, low completion rates, suboptimal coverage ranging from 2.4% to 73%, TB provider knowledge gaps, conflicting views on the importance of treatment, as well as the uncertainty about the duration of treatment, i.e. whether it should be administered for six months, nine months, 36 months or a lifetime [2, 3]. Shorter, safe regimens are crucial for increasing treatment completion rates and curbing the spread of active TB disease [2, 4]. These regimens include 3HR; a daily three-month (90 doses) TPT and 3HP; a weekly three-month (12 doses) TPT of combined rifapentine and isoniazid. The proposed three-month therapy to replace the standard IPT is 3HP as 3HR is offered in a few countries, only limited to children and adolescents aged less than 15 years and still administered over a long duration [2]. Additionally, randomized control trials demonstrated that participants taking 3HP had a threefold likelihood of treatment completion and improved outcomes compared to participants taking 6-9 month IPT [4]. The WHO, therefore, released the 2019 updated guidelines for the treatment of LTBI recommending the use of 3HP for people living with HIV (PLHIV) and contacts of TB cases of any age in place of the standardized daily self-administered IPT [1].

In alignment with the updated WHO guidelines, the National HIV/AIDS and TB Control Program under the Ministry of Health and Child Care (MoHCC) of Zimbabwe and partner non-governmental organizations (NGO) set the Tuberculosis national TB strategic plan of 2017-2020 (TB-NSP) which included the provision of 3HP in place of the current 6-month TPT at facility level [4]. The first phase of the program implementation (quarters one and two of 2020) was earmarked for 20 health facilities across five provinces in Zimbabwe [3]. However, the effectiveness and the success of the 3HP program might be a distant reality as there are operational challenges within the National HIV/AIDS and TB Control Program. Performance indicators are not being met including the sub-optimal performance of the current 6-month TPT [3]. Challenges include the shortage of skilled health workers, lack of capacity to recruit additional staff, lack of funding and training to support monitoring of TPT implementation exacerbated by stockouts of essential medication as a result of decades of crippling economic challenges [5]. Persistent IPT drug stockouts, for instance, resulted in the interruption of the TB prevention program [5]. As vital as the change from standard IPT to treatment by 3HP is, the aforementioned challenges could limit the successful implementation of the program.

Organizational readiness (OR) is vital and a necessary precursor to ensure successful implementation of the 3HP program in Zimbabwe. OR is defined as an implementation strategy to indicate the extent to which an organisation is psychologically and behaviourally prepared to implement a new program [6]. For example, an organization has to be willing and capable to implement a program for an effective outcome [6]. Studies indicate that about 50% of programs fail due to poor organisational readiness prior program implementation yet there is a dearth in the literature to indicate organizational readiness to implement the 3HP program among high burden, low-income TB settings such as Zimbabwe [7]. Such a gap in knowledge affects the optimum response to the TB preventive efforts of the country. An example highlighting the need to assess readiness before program implementation is the low utilisation rates of the GeneXpert innovation to improve TB screening in Zimbabwe varying from 50.4% to 63.5% despite its proven cost-effectiveness [5]. Similarly, piloting 3HP in facilities in Zimbabwe without assessing OR, its perceived barriers, and facilitators by staff collectively could present the failure to realize the real impact of 3HP. Therefore, determining OR is key to identify bottlenecks as perceived by the organization’s staff that could impede efforts to scale-up and to develop recommendations for better implementation.

The outcomes of this study indicated the readiness to implement 3HP and possible influencing factors within the selected facilities in Harare and Bulawayo Metropolitan provinces. By using this method, we hope the quantitative measurement strengthened by the lived qualitative experiences will identify operational metric challenges of the facilities useful to the National HIV/AIDS and TB Control Program and address issues that might otherwise escalate into obstacles that could impede successful program implementation [6]. This is critical because 3HP is a key TB prevention strategy to offset prior suboptimal IPT implementation efforts [3].

To date, no organizational readiness study for the 3HP program has been conducted in the sub-Saharan region, including Zimbabwe. The study will contribute towards the theoretical knowledge of implementation readiness before introducing 3HP in facilities and also expand on the lack of consensus on the measurement of readiness. Hopefully, the study will demonstrate the importance of considering readiness at the early stages of program implementation as it serves as a proximal indicator to successful, quality program implementation without undesired consequences. This will encourage monitoring and evaluation of the program at scale-up as well as tightening of potential loopholes in the implementation. Additionally, lessons learnt from the study can be adapted to facilities in the other provinces and later national level to give a more generalizable picture of 3HP implementation.

### Conceptualization of Organizational Readiness

In this study, elements in Weiner’s (2009) model attempts to understand the collective efforts or behaviour of members in an organization which lead to organizational readiness [6]. In the model, contextual factors, task demands, resource availability, and change valence are mediating elements of organizational readiness as a measure [6]. Change efficacy and change commitment are direct elements affecting organizational readiness [6]. Weiner highlights the importance of the interrelatedness of these factors that may affect readiness to implement a new program. For example, when members are committed, it leads the capability or strife to conduct the program implementation [6]. Change efficacy is based on three enabling factors, including task demands, resource availability and contextual factors of the organization [6] (S1 Figure).

**Fig 1.**
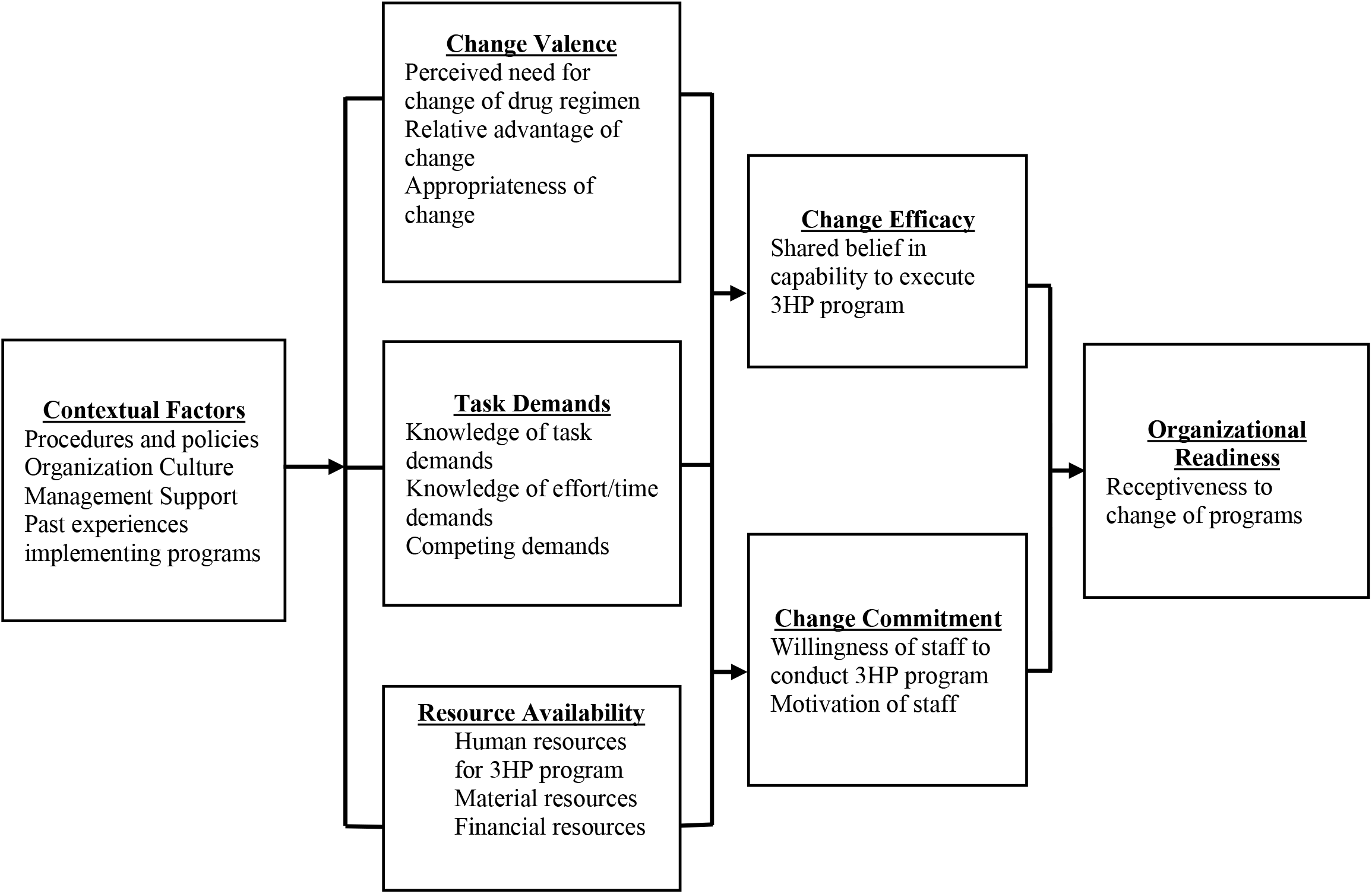
Weiner’s model for organizational readiness.

## Materials and Methods

### Study Design

This was a convergent, parallel mixed-methods study. Quantitative and qualitative data were collected in the same phase of the research process to measure organizational readiness and to explore barriers and facilitators that may affect readiness to implement 3HP in the four health facilities in Zimbabwe. The elements were analyzed independently, and the findings were triangulated and interpreted together. The purpose of this approach was to illustrate quantitative results with qualitative findings, synthesize complementary results for a more complete in-depth understanding of readiness and to compare multiple levels within the health care system [23].

### Study setting, population, and sampling

The National HIV/AIDS and TB Control Program in collaboration with the IMPAACT4TB program aims to pilot 3HP in twenty public health facilities across five provinces in Zimbabwe. In this study, data was collected from four health facilities across Harare and Bulawayo Metropolitan provinces. The facilities in Harare Metropolitan included Highfields polyclinic and Glen Norah Satellite clinic while in Bulawayo Metropolitan, the facilities included were Pumula and Northern Suburbs clinics. The four facilities were purposively selected based on physical location, similar facility sizes with respect to high out-patient volumes and same access to technical support. Additionally, Harare and Bulawayo Metropolitan were the only provinces that had infectious disease hospitals that decentralized TB prevention programs to 90% of their clinics, and hence it was expected that relatively higher out-patient volumes would access the 3HP intervention here [22].

The target population for this study were facility level staff (health facility managers or more commonly referred to as charge nurses, TB nurses and primary care practitioners) that had worked on the frontline and will directly implement the 3HP program. In addition, key informants such as a subset of the health facility staff (Charge nurses, TB nurses and primary care practitioners) as well as non-frontline personnel within the implementation support structure of the facilities (National level TB program managers inclusive of TB supply chain manager, the respective provincial-level TB coordinators from the National HIV/AIDS and TB Control Programs; and TB program managers from implementing and funding partners).

The study purposively selected 15 KII participants involved in the program that will implement 3HP in facilities. Concurrently, at each facility level, there was a maximum of 5 staff members involved in TB programs [5] and therefore we administered questionnaires to all 20 participants available at the four facilities. As per Weiner’s theory on organizational model guiding the study, readiness is measured at the supra-level and not at individual level [6]. Therefore, the study aggregated individual level data to represent shared perceptions within facilities.

### Procedures

Data was collected simultaneously among various target groups from February and ended in March 2020. Information sessions were held individually with the participants to inform them about the intervention, the aim, objectives and importance of the study. Thereafter, facility level staff (Charge nurses, TB nurses and primary care practitioners) answered hard-copy self-administered organizational readiness questionnaires to measure the level of readiness in implementing health facilities. Concurrently, open-ended semi-structured interview guides were used to understand the barriers and facilitators on the readiness with each of the fourteen (14/15) key informants lasting approximately 20-30 minutes. Written consent for audio recording of interviews and willingness to participate in the study was obtained from each participant. Both the qualitative and quantitative study tools had been pre-tested by the PI in Bulawayo Metropolitan before the main study and there was the need to modify the wording in the instruments for the participants’ improved understanding. Changes were channelled into the main study. The interviews were conducted privately by the PI in three of the main local languages namely English, Shona and Ndebele at the participant workplaces over a period of 5 days in Bulawayo and another 5 days in Harare. Data were collected as audio recordings and field notes. The audio recordings were obtained to allow the investigator enough time to listen and transcribe interviewees’ responses accurately. The transcripts were returned to participants for comment and/or correction. Field notes including descriptive and reflective information were captured in a diary by the investigator during or after interviews to ensure richness in the qualitative findings reported, interpreted and discussed.

### Measures

The questionnaire used 5 point-Likert scale responses that comprised of eight sections. Section A consisted of demographic characteristics. Section B-E supported an ordinal level of measurement. Section H was the additional comments section consisting of open-ended questions. With regards to section B-E, the Likert technique consisted of a series of statements to which one responded using a scale of possible answers ; 5-Strongly agree, 4-Agree, 3-Neutral, 2-Disagree and 1-Strongly Disagree. The questionnaire also included open-ended questions about perceived limitations and recommendations. Given that literature states that there is no standardized organizational readiness questionnaire, the structured questionnaire was developed from the constructs of the Weiner Organizational readiness model [6]. Cronbach’s alpha was used to measure the internal consistency (reliability) of the subscales and the composite scale [23]. The overall Cronbach alpha of the readiness scale adopted was 0.92, a score above the threshold of 0.70. While resource availability had a low reliability score of 0.50, change commitment, efficacy, valence, task demands and contextual factors to implement 3HP in the facilities all had reliability scores of 0.76, 0.84, 0.93, 0.73 and 0.82 respectively (S2 Table).

### Data Analysis

Data from the hard-copy questionnaires was captured using REDcap by both the PI and the data collector and exported into STATA 16 for statistical analysis. Cross-tabulations were performed in STATA to describe the characteristics of the facilities and socio-demographics of staff. Results were reported as frequencies and percentages for categorical data and means (standard deviation) or medians (interquartile range) for numerical data. Responses from the organizational readiness questionnaire which used Likert scale responses to measure the degree of agreeableness to statements were presented descriptively using median scores due to non-normally distributed sub-scales, based on the Shapiro-Wilk test. Subscale median scores > 3.3 indicated areas of strength which translate to the idea of “readiness” in that the respondents generally agree that the facility has the attribute in a given subscale. Conversely, scores < 3.3 show areas of weakness and need attention prior to change efforts. The Kruskal-Wallis rank test was used to test differences within and between the facilities. The significance level was set at p-value <0.05.

All interview data were combined with the responses to the open-ended questions in the questionnaire. The data were analysed as outlined by the framework method reported by Gale et al [25].Once all the data was coded using the analytical framework, data was manually summarized in a matrix through the charting process using Microsoft excel. The matrix compromised of one row per participant and one column per code. Data was abstracted from transcripts for each participant and theme, summarized using verbatim words and inserted into the corresponding cell in the matrix. The study found MAXQDA software to be invaluable at this stage for quick and easy retrieval of indexed data for specific themes within each transcript. Characteristics of KII participants including gender, duration in current position, program area and level of organization were retrieved and presented.

## Results

The survey participants (S1 Table 1) included nurses (73.7%, n=14), charge nurses (15.8%, n=3) and primary care practitioners (10.5%, n=2). There were more females (84.2%, n=16) as compared to the males (15.8%, n=3). Majority of facility 1 respondents were aged above 50 years (80.0%, n=4), while the majority of the respondents in facility 3 were aged between 40 – 50 years (60.0%, n=3) and similarly in facility 4, most of the respondents were aged between 40 – 50 years (50.0%, n=2). There were no participants (0%, n=0) who had provided TB services for 6 – 11 months however most had provided TB services for more than 2 years (79%, n= 15) with 100% of the facility 3 respondents having provided more than 2 years of TB service. Between the two provinces, Bulawayo Metropolitan had slightly more respondents in the facilities (52.6%, n=10) compared to Harare Metropolitan (47.4%, n= 7) (S1 Table).

**Fig 2.**
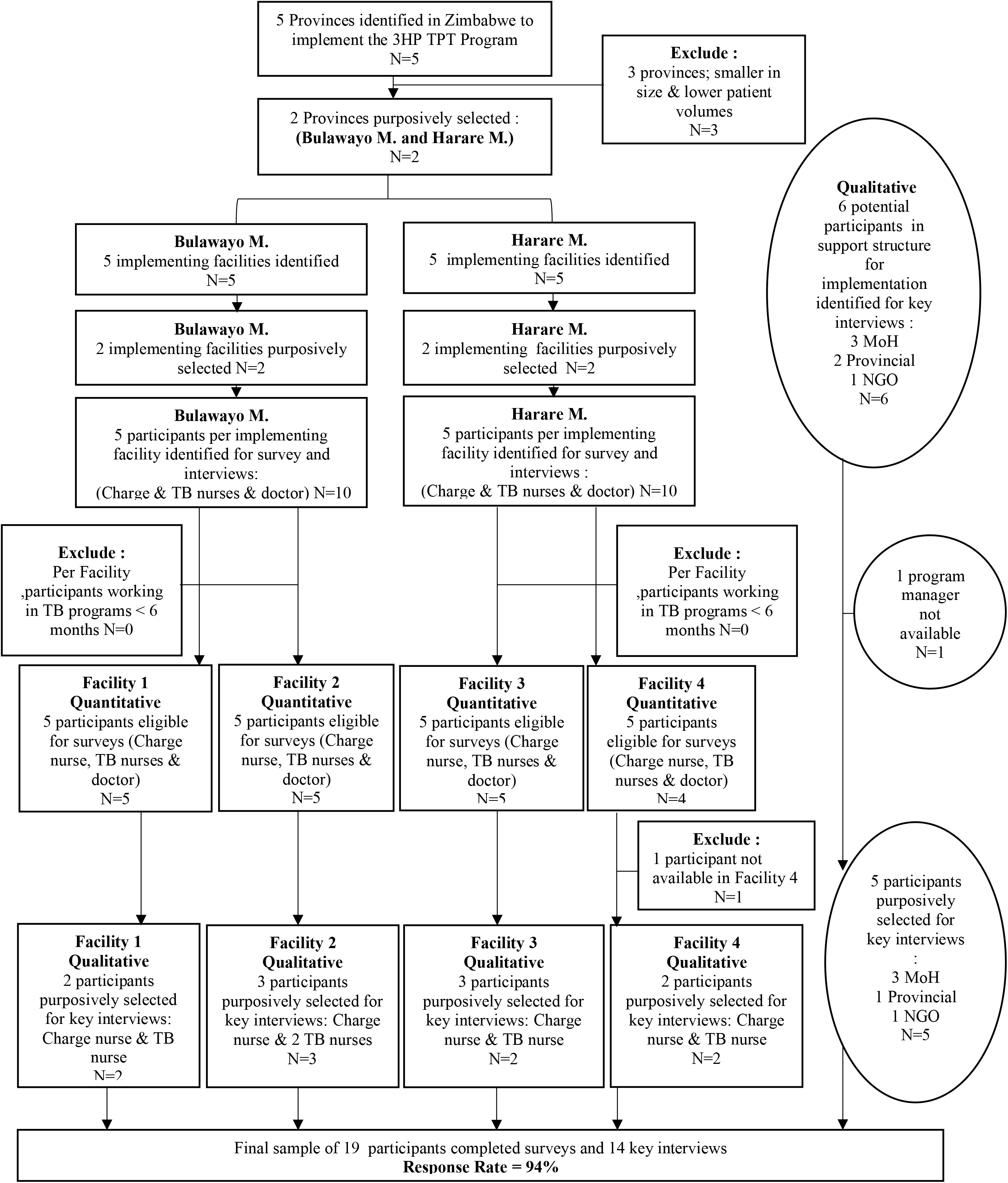
Flow diagram of selection of study participants.

### Level of organizational readiness in the four facilities

The overall OR median score across the four facilities was 3.8 (IQR 3.3 – 4.1) above 3.3, the chosen cut-off point for an acceptable level of readiness (S2 Table).

At the individual facility level, a total of three of the four facilities had median scores above the readiness cut-off point. Facility 4 within Harare Metropolitan province was below the cut-off of 3.3 as it had an OR median score of 3.2 (IQR 2.7 – 3.3). Low median scores were presented in the facility’s contextual factors 2.5(IQR 2.0 – 3.3), task demands 2.6 (IQR 2.3 – 2.9) and resource availability 2.1(IQR 1.5 – 2.5). The change commitment 3.7 (IQR 3.1 – 4.2), change efficacy 3.3 (IQR 2.8 – 3.6) and change valence scores 4.2 (IQR 3.3 – 4.8) of facility 4 were above the cut-off. Alternatively, Facility 3 within Harare province had the highest OR median score of 4.0 (IQR 3.8 – 4.2) although it presented a low resource availability median score of 3.0 (IQR 2.8-3.3).Consequently, Bulawayo Metropolitan’s facility 1 had a high level of readiness with an OR median score of 3.9 (IQR 3.9 – 4.0). The facility presented high median scores in the change commitment, efficacy, valence, contextual factors, task demands. Nevertheless, facility 1 presented a low median score of 2.8 (IQR 2.5 – 3.3) in the resource availability. Facility 2 in Bulawayo Metropolitan province had an acceptable OR median score of 3.6 (IQR 3.4 – 4.1) however a low resource availability median score of 2.8(IQR 2.5 – 3.3).

Significant differences were noted in the overall OR scores between the four facilities (p=0.039) (S2 Table 2). Within Harare province, significant differences were identified between facility 3 and 4 (p=0.016) wherein facility 3 readiness score was the highest compared to facility 4 which presented the lowest score below the threshold. Although there were no statistically significant differences in the change commitment, change efficacy, valence and task demands scores within provinces and between the four facilities across the provinces; significant differences were identified in the contextual factors between facility 3 and facility 4 within the Harare province (p= 0.016) and in the contextual factors between all of the four facilities (p=0.011). Marginal significant differences were found in the resource availability scores between the two Harare facilities (p = 0.048) (S2 Table).

**Table 1:** Demographic characteristics of participants in the implementing health facilities.

| Characteristics |  | Total<br>N(%) | Facility 1<br>n (%) | Facility 2<br>n (%) | Facility 3<br>n (%) | Facility 4<br>n (%) |
| --- | --- | --- | --- | --- | --- | --- |
| Sex |  |  |  |  |  |  |
|  | Female | 16(84.2) | 4(80.0) | 5(100.0) | 5(100.0) | 2(50.0) |
|  | Male | 3(15.8) | 1(20.0) | 0(0.0) | 0(0.0) | 2(50.0) |
| Age Group |  |  |  |  |  |  |
|  | 18 - 29 | 2(10.5) | 0(0.0) | 0(0.0) | 1(20.0) | 1(25.0) |
|  | 30 – 39 | 2(10.5) | 1(20.0) | 1(20.0) | 0(0.0) | 0(0.0) |
|  | 40 – 50 | 7(36.8) | 0(0.0) | 2(40.0) | 3(60.0) | 2(50.0) |
|  | Above 50 | 8(42.1) | 4(80.0) | 2(40.0) | 1(20.0) | 1(25.0) |
| Role In Facility |  |  |  |  |  |  |
|  | Charge nurse (Facility manager) | 3(15.8) | 1(20.0) | 1(20.0) | 1(20.0) | 0(0.0) |
|  | TB Nurse | 14(73.7) | 4(80.0) | 4(80.0) | 4(80.0) | 2(50.0) |
|  | Primary care practitioner | 2(10.5) | 0(0.0) | 0(0.0) | 0(0.0) | 2(50.0) |
| Years Of Providing TB Services |  |  |  |  |  |  |
|  | 6 – 11 months | 0(0.0) | 0(0.0) | 0(0.0) | 0(0.0) | 0(0.0) |
|  | 1 year to 2 years | 3(15.8) | 1(20.0) | 1(20.0) | 0(0.0) | 1(25.0) |
|  | More than 2 years | 15(79.0) | 3(60.0) | 4(80.0) | 5(100.0) | 3(75.0) |
| Province |  |  |  |  |  |  |
|  | Harare Metropolitan | 7(47.4) | 0(0.0) | 0(0.0) | 5(100.0) | 4(100.0) |
|  | Bulawayo Metropolitan | 10(52.6) | 5(100.0) | 5(100.0) | 0(0.0) | 0(0.0) |

**Table 2.**
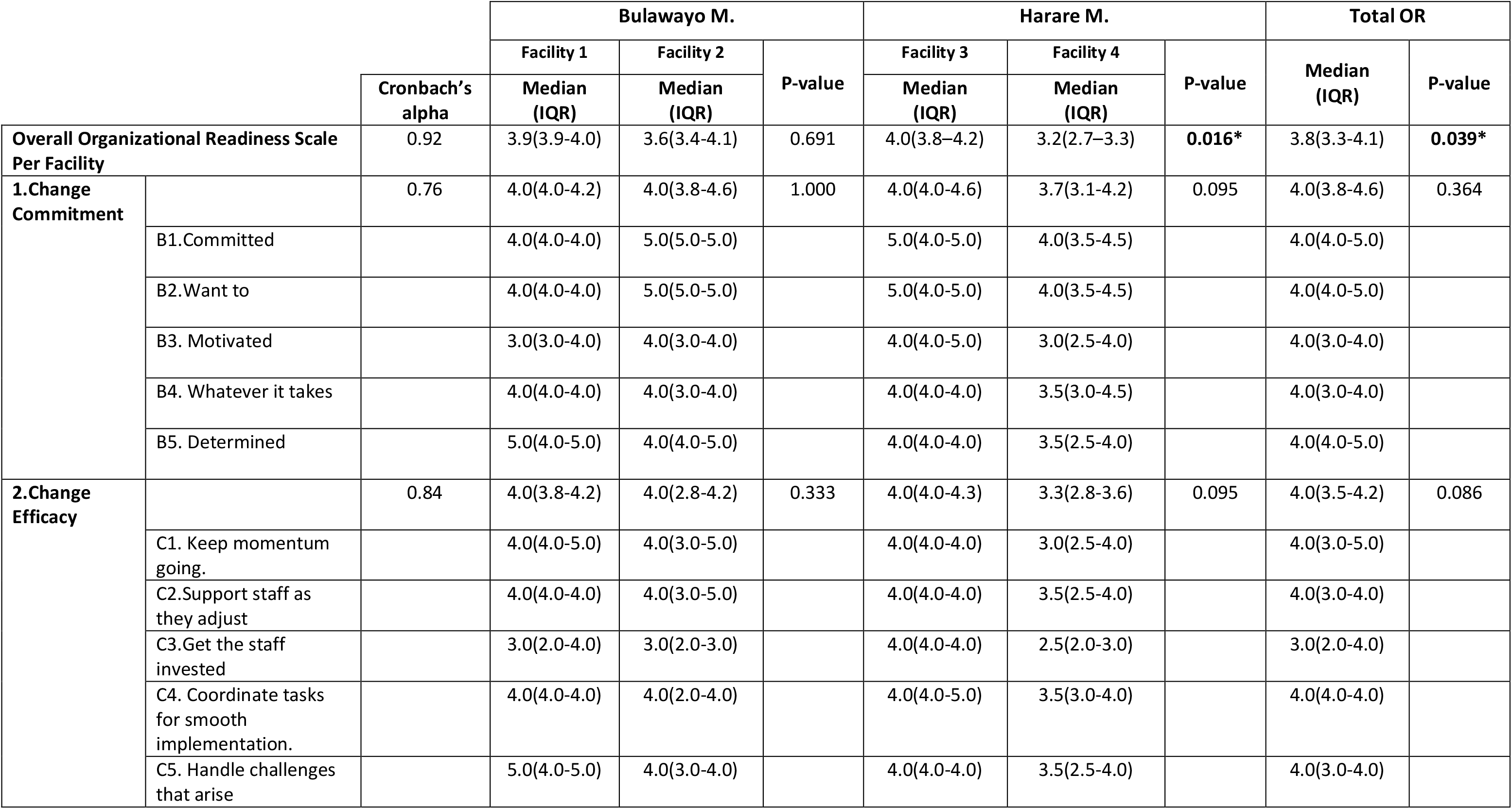
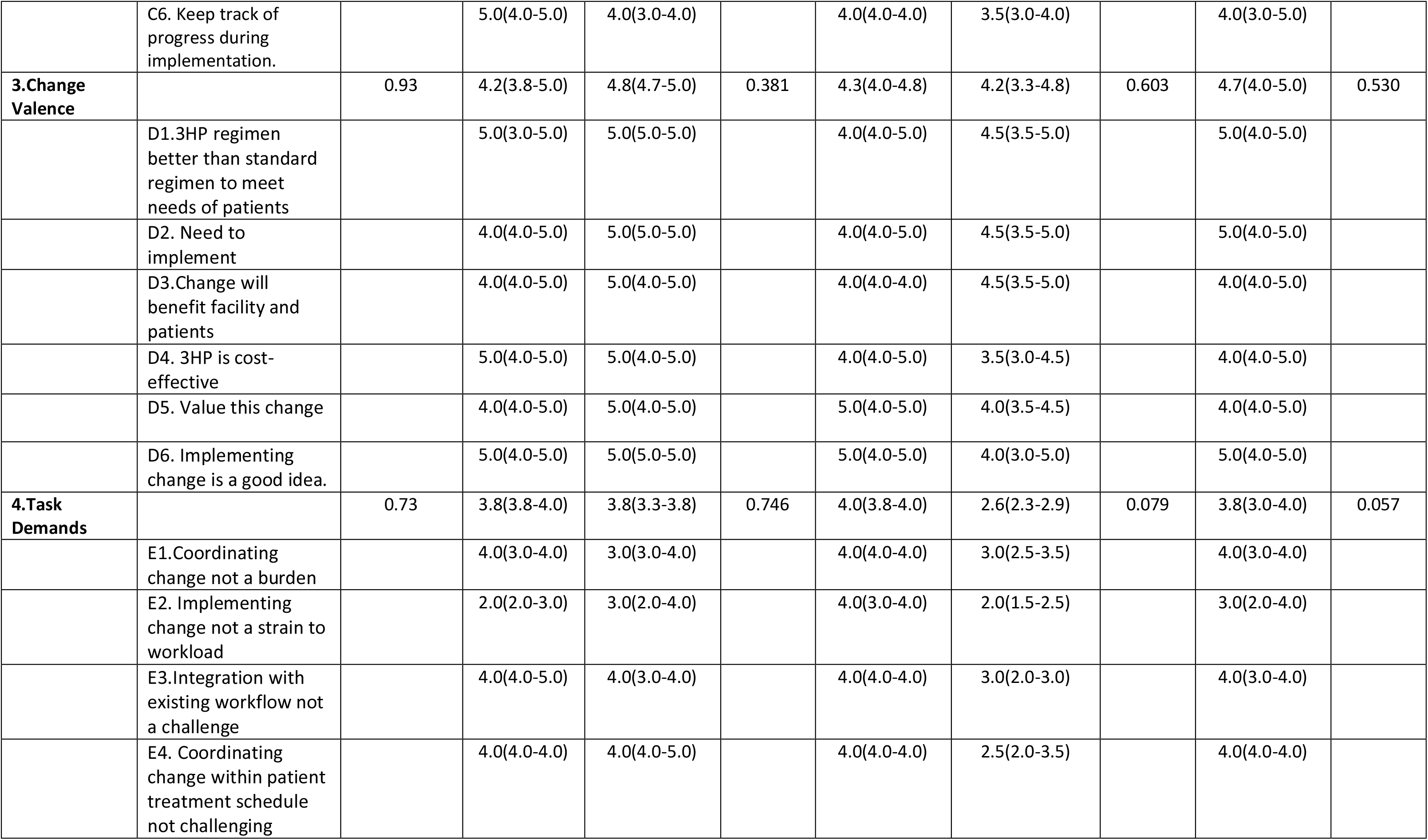
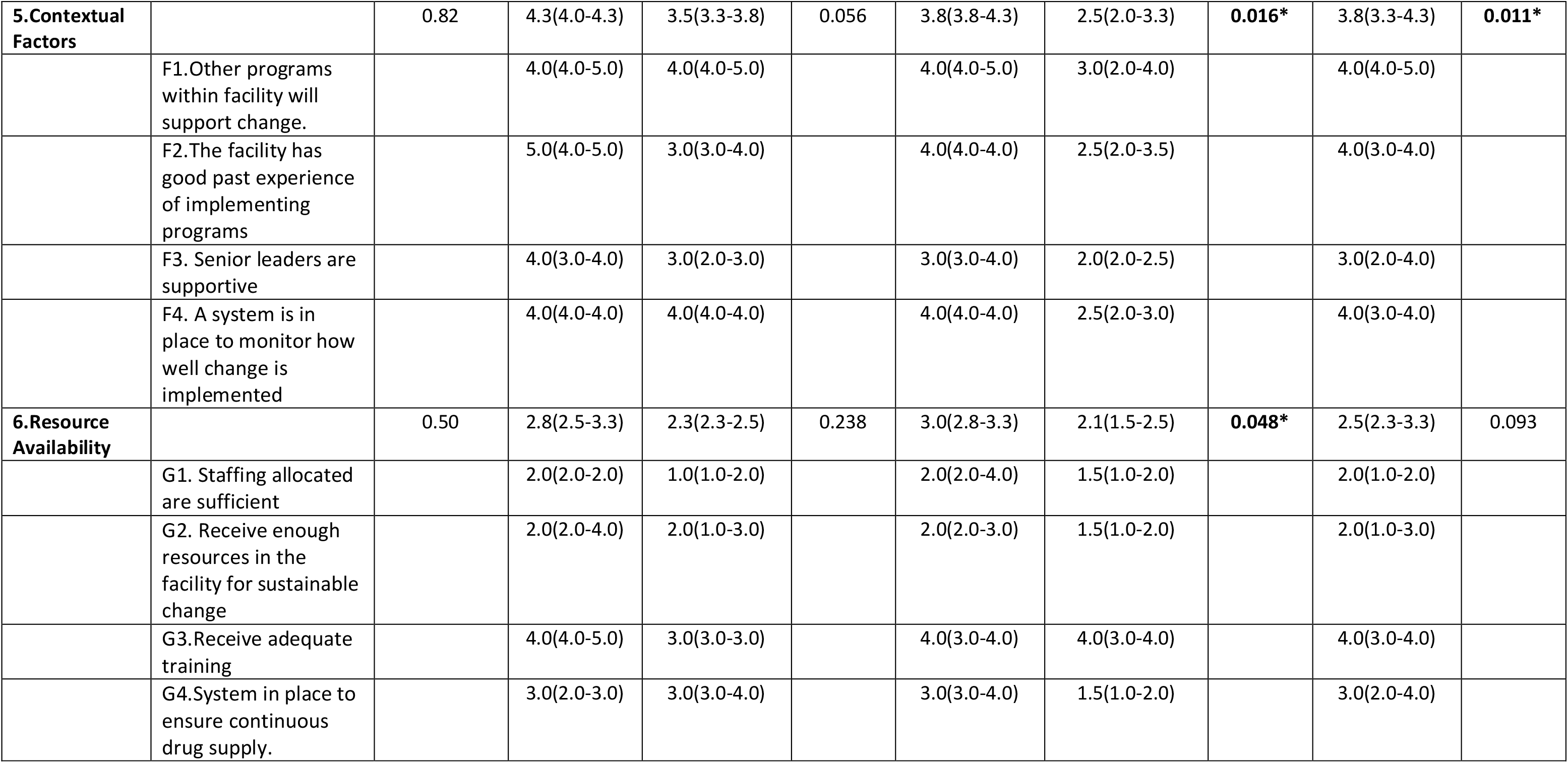
Organizational Readiness scale and subscale scores.

### Perceived Barriers and Facilitators

Six (n=6) TB nurses and three (n=3) charge nurses were among those interviewed at facility level. Additionally, five (n=5) program managers from the implementation support structure of the facilities (National, provincial and NGO levels) were interviewed. Four participants were male while ten were female. Work experience ranged from 1 to 17 years.

### Change Commitment

#### Facilitators

The managers in the TB program believed that there was an overall stakeholder buy-in and willingness to commit to the program change. A program manager with 3 years’ experience believed that *“it is worthwhile to change to the 3HP*.*”* Similarly, most facility staff were optimistic and they expressed willingness for implementation. A facility 1 TB nurse reported that

> *“We are positive, and we are willing to adapt to the program and implement it*.*”*

Managers also generally reported excitement through demand generation for 3HP created by organizations and civil societies and patients. *“We have been working with a lot of community support organizations and civil societies. They have also been generating demand in communities and so yah I can definitely say there is a lot of excitement… So, this is not just health care workers pushing it to patients but actually patients coming in to ask to receive this therapy in place to the current one they are on, “* explained a TB program manager from a partnering NGO.

#### Barriers

Commitment was indicated to be temporary and contingent on factors including unsatisfactory remuneration and an inconsistent supply chain management in the facilities. For example, a program manager stated that

> *“The shortage of medicines can draw back the motivation of health care workers*.*”*

Facility 1 staff reported burnouts due to workload therefore rewards were necessary to motivate them. A charge nurse reported that

> *“When it comes to burnouts, [pause] they will be complaining that they are tired and they will not be motivated…The incentives motivate them*.*”*

Although most of the facilities indicated conditional commitment, facility 4 respondents were not willing to implement the program due to their perceived on-going facility issues. *“Considering we have our own challenges already yeah I don’t think people will quickly embrace it, “* explained a TB nurse.

### Change Efficacy

#### Facilitators

The management level believed that similar to the HIV program, the nurses could easily lead the implementation and oversee the program change in the facilities. A program manager explained that:

> *“…our HIV program is nurse-led, happening at the lowest level of facilities, so they are very well capable to implement this program*.*”*

Management believed that the change was going to be a *“clear process”* (IDI 1) while facility 2 staff stated that the program was going to be straightforward given that they were going to be trained and given guidelines. A TB nurse described that

> *“where there are guidelines, its easy*…*the person should have time to read over and understand what they mean. As soon as they grasp them, they are easy to follow. The guidelines actually help and are easy to understand*.*”*

In a bid to increase acceptance and accountability, management aimed to sensitize “90 to 100%” of the staff in facilities.

#### Barriers

Both levels of participants reported negative staff attitudes.

> *“… we had negative attitudes from the service providers and then the patients themselves… knowing the possibilities of the adverse events*.*”* (IDI 1).
>
> *“The problem is that most of the nurses have attitude towards those patients. I am sorry to say (uhm) but that is very natural. Attitude towards those TB patients with the fear that they will catch TB in the process of nursing them… “* (IDI 12).

The staff also reported a lack of confidence in the momentum. A TB nurse described that *“…the kickstart will be okay but on the way you find some problems cropping up…like there can be no medicines for some time*.*”* Additonally, there remains a challenge of misinterpretation of the Ministry guidelines. A TB medicines supply chain manager explained that

> *“…we realized that the staff were initiating people on the current regimen yet they were not eligible*…*so the staff had everything and the eligibility criteria but their interpretation of that eligibility criteria was different and so those are some of the things that we need to put into consideration*.*”*

Both the management and facility level staff also stated that the informal nature of the communication within the program caused a communication gap. A MoHCC manager acknowledged explained that

> *“We write memos from national to province, province to district, district to facility. If there is break in the communication channel, it is possible to get to a facility and they tell you that they have not received that communication, so they are not doing anything about it*.*”*

Participants reported that program coordination is affected by the flexi-hour system where nurses rotate and only work two days a week. Nurses are transferred for the next rotation immediately after familiarizing themselves with the day-to-day management of the program, and the next person replacing has to familiarize with that work station again. A TB nurse explained that

> *“Now the rotations that are there, the TB nurses or the OI nurses that are there are so used to the routine. Everything is smooth sailing and then you have to be changed*… *I’m said to go to maternity then somebody new has to come here to OI and start adapting to this whole thing. For a while that would be a challenge*.*”*

Lastly, systematic challenges including a poor monitoring and evaluation system was reported by all the respondents as there was said to be’…*a great need for management to work with facilities on the ground’* at the early stages of implementation to enable strengthened data management and reporting.

### Change Valence

#### Facilitators

A positive factor was the perceived program appropriateness which was the relevance and compatibility of the 3HP innovation to address problems within the existent program. A TB program manager stated that *“we do believe a lot of issues from the previous regimen, IPT will be addressed…”* Similarly, the change effort was perceived as a need and was described to be timely as to alleviate medicine stockouts that facilities were experiencing. *“The timing is right…we are running short of the current IPT medicines,”* explained a TB nurse.

Additionally, all facility staff expressed that the program would be regarded as a priority. A charge nurse echoed this as follows,

> *“TB is one of the priorities in any council clinic so I believe if 3HP is a prevention strategy as part of TB programs, it would be a priority here*.*”*

The staff further discussed the relative advantages the program may offer such as lower costs and risk of mortality for their clients. A TB nurse and focal person believed that *“…it actually brings with it a lot of relief, to the community in the sense that it provides low cost strategy in the long run. Patients are not required to continue coming to the clinic because treatment duration will be cut down which reduces the frequency of visits so it is less transport cost for them*.*”* Facility staff also believed that the program would be of beneficial value to them and their health indirectly as they feel the treatment may reduce the risk of TB transmission within their workspace.

#### Barriers

The recurring perception was that if patients were not interested nor valued the regimen, the program implementation would be negatively affected regardless of high valence among facility staff. This was a focal point for seven of the fourteen participants, and examples of their responses included:

> *“Yes, we are saying 3HP will produce good results but the challenge comes on clients*… *clients are already complaining of taking a lot of pills”* (IDI 14).
>
> *“The 3HP pill burden is huge*…*So for us we might be happy that it is a short regimen but the burden then comes to the patient or the recipient of care. They are the ones who are going to decide whether they are willing to take 10, 13 tablets once a week at once or they want to take 1 or 2 tablets every day for 6 months*.*”* (IDI 2).

The trade-off between a higher pill burden over a shorter period of time versus lower pill burden over a long period of time could affect the patient value of the program change.

### Task Demands

#### Facilitators

Managers reported that staff in the facilities will be provided with the materials and tools necessary to guide them with little effort required. A MoHCC program manager explained that

> *“ A lot of printed flow charts will be stuck on the walls of the facilities*…*even without opening the book but by just looking on the wall, they can see any SOP and they will be easily guided*.*”*

A provincial manager anticipated smooth integration of programs as rendered previously and stated that *“we have implemented programs in the past very well…the IPT was already integrated so we would think we will have no problems integrating this TPT into the program*.*”* Overall, management expressed positive insights in contrast to facility.

#### Barriers

Regardless of the perceived basic effort reported by the managers, there was an anticipated challenge of increased workload, specifically registers with the introduction of the 3HP program amongst the staff in all of the facilities. A charge nurse explained that “complex registers” strained their workload and reported that

> *“Each program comes with new registers*… *we have so many registers besides our work to enter*…*we have to fill this register, now I go to the EPI program there are registers, you go to the OI program - a heap of them and now the TPT [laughter]*.*That’s the only challenge we can really have with the work load*…*”*

Although some facilities reported that the increased workload was due to facility staff shortages, a MoHCC program manager stated that the workload should not be a challenge as staff were expected to *“synchronize their activities, to harmonize their service provisions and provide services under a one stop shop approach”*

The second barrier included time constraints within the staff schedules. A charge nurse reported that there was an inability to complete all their tasks and stated that they “…*cannot help all of the patients in a day, the ratio of all patients to health personnel is hectic*.*”*

### Contextual Factors

#### Facilitators

Managers stressed that the importance of the initial implementation phase was for management to show support to their facilities which in turn will allow for staff to follow suit and support their TPT patients. A MoHCC program manager echoed and explained this

> *“ … we will continue to visit facilities and make sure that whenever they come across patients, they will support them the same as we get support. That is the importance of this initial phase*.*”*

Moreover, management and staff both also believed that facilities are accustomed to research and innovation which will facilitate change implementation. A charge nurse explained that

> *“People are not resistant to change. There are a lot of programs that are coming, and they are dynamic and there is need to move we cannot stay in one place*.*”*

#### Barriers

Support and managerial oversight is inconsistent in facilities affecting facility staff. Support of staff in terms of their wellbeing was no longer made available as in the past which exposes them to infectious TB patients. They explain that they are *“exposed waiting to get sick*.*”* A TB nurse and focal person described that managerial oversight is inconsistent affecting facility staff.

> *“Visit us periodically. Not just introducing a program and go [laughing]…In the [name of organization] program it is also a TB program, we only did a workshop last year. They are not following up on us*.*”*

Facilities also stated that they have experienced negative encounters such as adverse effects on patients while implementing TPT programs which the managers reported that they are aware it can impact staff confidence. A TB nurse recounts her experience

> *“What happened here at the clinic*…*there are these bad side effect that happened to our patients eeeh [shakes head]* …*one of my colleagues, we were together in the room when a patient developed [pause], it was like a syndrome or something like that. The patient is in a wheelchair till today, whenever they come to clinic, we feel bad. What if it happens to me that I am the one with two or more that are put in wheelchairs?”*

The managers believed that the whole organization *“…has not been performing well in the last few years”* regarding TPT and the culture of slow program implementation in the facilities could affect this program change. A nurse gave an example;

> *“At times they take long because of the availability of the drug for example introducing [name]. It was introduced in 2015, we only started giving it last year [laughs] as in 2019. Four years later*.*”*

### Resource Availability

#### Facilitators

Managers at national level reported that efforts at central level were made for the program’s smooth logistic flow in facilities.

> *“We look at our plan holistically and based on what we can get, we start off by initiating 10% of our clients and monitor the progress. That is how we see that our supply and programming match. This reduces ruptures within the supply in our programs…there is also a mechanism in place where facilities can place emergency orders from the central stores and depending on the province they are in, there is a turnaround time of about two weeks”* (IDI 3)

The NGO program manager also explained the importance of partnerships between organizations and the Ministry for the program’s funding.

> *“We also made sure to choose sites supported by PEPFAR to ensure sustainability of this work. One; because these are supported in terms of staffing, and two; TPT scale up is a priority with PEPFAR”*

#### Barriers

Despite the efforts to improve the logistic flow at national level, provincial managers and staff stressed that the irregular supplies of the 6-month IPT program affected their perception of the change effort.

> *“…there were erratic supplies of late*…*the logistic flow was not good. You find that drugs are plenty in one province and in another there is nothing*.*”*

Staff reported recurring human resources shortages at facility level whereas management stated that *“there is high turnover of the staff…they leave the facilities after being trained*.*”*. Moreover, staff reported a lack of incentives and irregular remuneration due to management not being favourable of the concept. Additionally, the NGO program manager reported that production capacity and 3HP pricing were external barriers to the supply chain beyond control.

> *“ The supplier’s capacity did not turn out to be what they had initially committed [pause] so they could not commit to the volumes that our country needed*… *another thing was that the drug price … these are things that delayed rollout at country level*.*”*

It was further stated by the manager that *“…we anticipate further delays now with the COVID-19 pandemic*.*”* Nevertheless, provision for sustainable funding after the initial implementation phase remains unclear as the program heavily relies on partnering and actively present NGOs within the facilities. An application for funding towards the scale-up of the program was still in the development stages. The partnering NGO recommended that *“clear commitment for funding would enable the program to include all eligible patients without adjustment of the guidelines*…*”* (S3 Fig).

**Fig 3.**
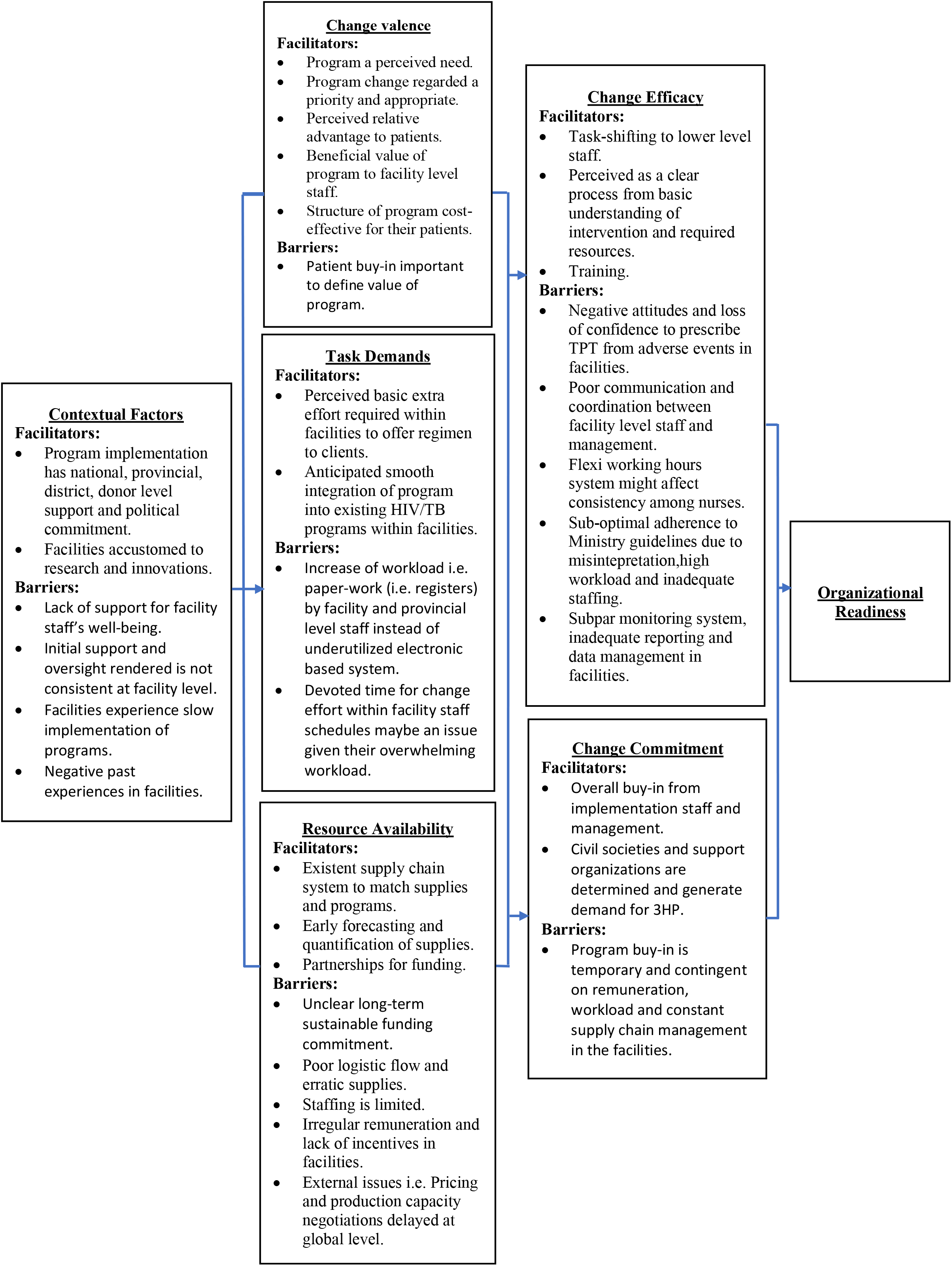
Summary of key findings.

## DISCUSSION

The findings from this study identified fundamental facilitators and barriers which will prompt further planning and improvement to the 3HP program prior to implementation in the four facilities. The improvements could influence implementation effectiveness.

### Level of Organizational Readiness to Implement

Three of the four facilities (facility 1,2 and 3) welcomed and saw a need to implement the intervention. Our findings are supported by the measurements of readiness scores per facility which were above the threshold. We believe the high scores demonstrated that staff in the three facilities were most likely to collectively commit and demonstrate a more consistent, high quality use of the intervention [6]. The results led to a similar conclusion in Shaw et al’s study where high readiness scores reflected the degree of commitment and efficacy among facilities towards implementing a telemedicine behavioural program [15]. This is consistent with a South African study that used readiness scores to indicate that four of five schools collectively valued a combined HIV and nutrition intervention highly enough to commit to its implementation. Similar to our study, the two mixed methods studies used adapted context-specific tools to measure readiness [6, 15]. Although this is widely accepted, an apparent limitation is that the tools were adapted from contexts outside of the clinical or health fields due to limited healthcare readiness studies [31]. It is worthy to further note that literature indicated a paucity of studies assessing the level of readiness to implement 3HP in the Zimbabwean context and the few studies on 3HP implementation were based in high-income countries such as Canada.

Our results also demonstrated that facility 4 of Harare Metropolitan province was not ready to implement 3HP compared to the other 3 facilities. Additionally, staff in the facility qualitatively indicated that *“… we have our own challenges already…I don’t think people will quickly embrace it*.*”* At this stage of understanding, Weiner’s theory suggests that low readiness reflects the need for additional support before implementation to curb the facility potentially resisting change, putting less effort, persevering less in the face of challenges, and exhibiting compliant intervention utilisation [6]. Our finding also showed that the low readiness in facility 4 was due to low contextual factors, task demands and resource availability. The result of our study is then compared to Shaw et al’s study which found non-readiness in facilities due to a lack of commitment [15] and due to contextual factors including leadership, fidelity, costs and partnerships in another review [32]. The difference in the studies is attributed to the main limitation of the inconsistency of constructs measured which may impact readiness to implement. For example, our study did not account for leadership whereas Rubenstein et al’s study used the CFIR framework which found the construct to be key in the non-readiness of facilities [33]. However, a review suggested that there is no gold standard in readiness studies as existing issues were tailored to specific studies, interventions and settings [34]. From this standpoint, we believe our study is the first to contribute important findings specific to the 3HP intervention and possibly other TB program readiness studies.

Our findings on the difference in facility scores at least hinted that each facility had specific underlying conditions for readiness to implement 3HP. For example, one Harare Metropolitan province facility had the highest readiness score while the other was not ready and had the lowest score among all of the facilities. The difference between the two facilities was due to varying contextual factors and the availability of resources which was supported by the qualitative findings as well. This is consistent with two previous studies whereby intervention sites differed in readiness due to varying resource availability and reported contextual conditions such as past experiences in implementing programs per the specific setting [33]. It is worth discussing that our study’s approach of including facility staff plus supplemental qualitative accounts from different levels of management provided more detail on the varying conditions that lead to readiness as compared to Rubenstein et al’s study that indicated a limitation of only including reflections from facility leaders as a representation of the facilities [33].

### Perceived Barriers and Facilitators of Organizational Readiness

The findings demonstrated that change commitment, efficacy and valence were key facilitators of readiness to implement 3HP. The key barriers emanating were task demands, contextual factors and resource availability.

### Change Commitment

Change commitment was high among all the facilities and the finding was qualitatively supported by the perception that there was overall buy-in from stakeholders. This result ties well with previous studies that indicated that shared interest and commitment between managers and staff were found to be important precursors for a successful intervention [19, 25,6]. Although staff in the study indicated that they were motivated, there was the possibility of temporary buy-in dependent on issues such as inconsistent staff remuneration, increased work load and an inconsistent supply chain in facilities. This is challenging and has led to frequent strikes in Zimbabwe among frustrated health care workers disrupting implementation of programs [35]. A similar pattern of findings were obtained in Zhang et al’s study suggesting that discrepancies between the current and desired conditions within an organization overshadowed the high change commitment for programs [26]. Notwithstanding, our study had limited time and resources therefore was conducted at one-point in time as compared to the aforementioned previous study that measured readiness at the pre-adoption, pre-implementation and post-implementation stages of the program [26].

### Change Efficacy

There was high collective capability (efficacy) scores which paralleled positive qualitative findings such as the availability of refresher trainings, task-shifting of services, the anticipated basic and clear process of the implementation in facilities. However when comparing our results to those of older studies, it was indicated that efficacy may potentially be affected by varying factors. Firstly, although staff may report high efficacy to perform tasks, it is possible that they may not be able to create meaningful change due to the inability to mobilize resources in an efficacious manner for the desired implementation effort [6]. Secondly, the efficacy to collectively perform tasks can be affected by commitment and vice versa. For instance, Keshni (2020) stated that readiness is likely to be high when members not only feel confident to perform tasks but also want to do so [14]. Similarly, we demonstrated that the lack of confidence to implement previous TPT was attributed to by negative staff attitudes. Therefore our findings contribute to the idea that efficacy is interrelated to commitment. Lastly, apparent systematic challenges such as task co-ordination and flexi-working hour system in the facilities may affect efficacy. In this case, our finding lends support to earlier studies which popularly explain that discrepancies in efficacy are due to the differentiated roles in facilities (i.e. nurses, charge nurses, administrators and medical directors) [21, 28]. Therefore for future work, it is worthy to note that some questions in readiness tools may have more of a clinical meaning for participants, for example in our study that were majorly nurses as compared to other studies that had mostly leaders who may have more of an administrative meaning [28].

### Change Valence

Measured scores were supported by recurring textual findings that indicated that staff collectively valued the program highly. Staff felt that the previous suboptimal TPT implementation would be alleviated by the benefits of 3HP implementation, considered it a core priority within all facilities and comprehended it enough to commit to its implementation. This is warranted by staff acknowledging that they were willing (commitment) to implement the program because change in programs was urgently needed for the well-being of their patients. Our findings therefore indicate the link between change valence and commitment to positively influence readiness to implement. Our study is in line with Weiner’s theory (2009) and Keshni et al’s study whereby change commitment was proposed as a major function of change valence, that is, if organizational members value the change then the they will want to implement the change [6,14]. However, the main limitation is that individual perceptions are given in each study instead of collective staff perceptions of the program value which may implicate consistent, cross-situational relationship between change valence and organizational readiness. Future studies need to indicate if change valence resulting from different individual reasons might be as potent in readiness outcomes as change valence resulting from commonly shared reasons. Additionally, although the study reported high valence, facilities stated the challenge of an oversight of patient input on the value of the 3HP program. In line with our findings, a study posited that patients are often omitted from the implementation planning phase yet their value of the intervention holds a considerable amount of weight with regards to the implementation outcomes and program performance [15]. The patients’ perceptions would reflect if change valence truly influences readiness to implement despite the acceptable overall change valence score.

### Task Demands

Our findings showed recurring challenges mostly in facility 4 despite staff having anticipated little effort to offer the new regimen and a smooth integration of program activities within existing HIV/TB programs. Barriers as described by the staff included increased workload in the form of paperwork instead of availing the underutilized electronic system. The increased workload was mostly attributed to a lack of human resources in the facility as staff recommended that increased human resources could offset the strained workload. Shea et al’s study findings were consistent with our findings and similarly recommended for the recruitment of more staff plus provision of additional support to improve readiness levels [28]. Another identified challenge was that staff may not have devoted time for the program within their overwhelmed schedule which might impede implementation. Keshni et al’s study explains that if the facility is not inclusive of implementation efforts in the staff schedules, slow implementation will occur [14]. Our study recommends that facilities should develop and implement work transition plans to develop continuity plans which would mitigate both barriers presented with regards to task demands.

### Contextual Factors

Our study quantitatively confirmed that the facilities had distinct, acceptable contextual conditions however the qualitative findings detailed a myriad of challenges particularly in facility 4. For instance, support in facilities was rendered by key stakeholders however it was inconsistent and a further lack of support for the staff’s wellbeing was reported which both led to their negative attitudes. Izudi revealed that TB staff commonly present negative attitudes due to fear of contraction of TB from their patients affecting their performance to carry out tasks [29]. The provision of routine testing for staff implementing the TB program in facilities was recommended by staff in our study. Another challenge was the slow implementation of programs, for example facilities reported 4 year delays after trainings to implement programs. Additionally, our results suggest that the high number of adverse events associated with TPT contributed to the negative past experiences in facilities. We believe these combined could result in staff not embracing the innovation and therefore could impede readiness just as Amatayakul concluded that negative experiences could deter facilities from reaching organizational readiness [30]. The difference in studies, however, is that our findings go beyond to show that negative past experiences negatively affect the staff’s value of the program and their capability to implement it. Therefore, our findings contribute further to readiness evidence in demonstrating that contextual factors affect valence and efficacy which are proximal factors for readiness. Nevertheless, our study was guided by Weiner’s framework as compared to Amatayakul’s study which is guided by the Consolidated Framework for Implementation Research (CFIR) with different constructs [3]. Although facilities faced multiple contextual barriers, the staff indicated that they are accustomed to research and its rapidly changing nature which may contribute towards acceptance of implementation efforts.

### Resource Availability

Resource availability was not at a sufficient level for the implementation effort to occur effectively which was supported by identified qualitative barriers. Recurring challenges were the shortage and the inequitable distribution of staff as observed between facilities 3 and 4. The finding may be warranted by the smaller number of staff that were available in facility 4 as compared to the other facilities in the study. It was recommended that facilities should allocate personnel to serve as implementation champions to facilitate the process as similarly suggested by Ochurub, Bussin, Goosen [19]. We further demonstrated the lack of staff incentives that negatively impacted willingness. It is generally accepted that incentives and rewards increase job satisfaction, teamwork, as well as help staff, feel equipped to undertake a new initiative [14]. Alternatively, Scaccia et al’s [25] findings argue that staff may be motivated enough to work with whatever resources are available. The difference in study contexts may have led to the difference in conclusions therefore we believe that contextual conditions may play a key role in facilitating implementation despite resource inadequacy. However, given poor contextual conditions in Zimbabwe’s health care system [36], our study argues for more planning and the actual provision of resources to enhance facilities’ chances of readiness to implement.

Managers explained that unforeseen, external issues occur such as 3HP pricing and production capacity delays exacerbated by the onset of the COVID-19 pandemic which cannot be resolved at the country level. This is broadly in line with a study that highlighted the unexpected disruption in the implementation of HIV programs in Sub-Saharan African countries due to priority shifts at the start of the pandemic [37]. It is worthwhile to note there were positive aspects identified by managers which may overcome unforeseen consequences such as the early forecasting and quantification of supplies; and fostered partnerships for funding for the first phase of implementation.

### Strengths and Limitations of the Study

The main limitation of the proposed study rests primarily on the sample size particularly the survey sample which may not be statistically representative however the aim was firstly to analyse at the facility, not the individual level and to strengthen understanding of the social processes involved in implementation therefore sample size is of less concern. Although the sample size was large enough to measure OR in the study, it restricted further additional statistical analysis. Notwithstanding, Weiner, suggests that OR measures should be descriptive and not evaluative (6). Moreover, the generalizability of the results may be limited to the four facilities that were operated by the same entities however as in implementation science, the key is to draw on lessons learned to inform policy and implementation approaches in similar contexts(6).

Although the ORIC tool was adapted to suit the study context and design, the resource availability subscale was slightly below the threshold Cronbach alpha level. However due to the shown acceptable overall Cronbach alpha scale indicating the tool’s high reliability in consistency, the subscale was retained. Another limitation is construct bias wherein the selected Weiner OR conceptual model to guide the study may have omitted other constructs that affect readiness including implementation climate (14). Additionally, constructs within the conceptual model overlap and are not always discrete. Future studies are required to improve OR models and their constructs. Nevertheless, the current study provides insight into readiness factors that relate to the implementation of a TB preventive therapy which has received little attention thus far.

The study employed mixed methods which provided several merits. The combination and triangulation of the quantitative and qualitative analysis methods facilitated understanding the complexity of organizational readiness to implement 3HP. The quantitative helped measure the level of readiness and its factors as a useful benchmark of the sites. The qualitative offered insight into the challenges and influences of readiness. The interviews were held until saturation was achieved which is important in exhaustively underscoring the reasons for the measured scores in facilities.

## Conclusion

In conclusion, three of the four facilities were ready to implement 3HP in Harare and Bulawayo Metropolitan province. Facility four of Harare Metropolitan province was not ready to implement due to low task demands, contextual and resource availability factors. Each facility had differing strengths and weaknesses to be deemed ready to implement the intervention, but these would likely not be revealed without triangulation of OR scores and interview data. Potential factors that could either facilitate or hinder effective implementation were identified to allow enhancements to be made to the intervention implementation strategy. Facilitators identified were change commitment, efficacy and valence which should be considered in developing the steps towards meeting acceptable readiness levels to implement 3HP in facilities. The barriers included were task demands, contextual factors and resource availability.

The study findings demonstrate the necessity for further research to assess OR prior to 3HP TPT implementation in health facilities. Considering the limitation to generalize our findings, the study should be scaled up in other high TB settings in Zimbabwe as well as the other high TB burden countries yet to implement 3HP. Although implementation of the TPT program change will be staggered into phases, our study assessed readiness at one point in time as opposed to continuous assessment. We recommend the periodic evaluation of readiness within health facilities for optimal delivery of the program to ensure successful implementation. It is worthwhile to note that conditions for readiness per facility can differ therefore OR is critical for efforts to carefully attune implementation according to the strengths and barriers present at each facility. Moreover, policymakers need to take into account that facilities with less readiness to implement may require more flexibility to reach the intended program merits.

## Data Availability

The data that support the findings of this study are available from the authors however restrictions apply to the availability of these data, which were used with the permission from the Bulawayo and Harare City Council Health Service Departments for this study, and so are not publicly available. Data are however available from the authors upon reasonable request and with the permission of the respective health service departments.

## List of Abbreviations

3HP: Three-month short-course TB Preventive Therapy Regimen
HIV: Human Immunodeficiency Virus
IMPAACT4TB: Increasing Market and Public Health outcomes through scaling up Affordable Access models of short Course preventive Therapy for TB
INH: Isoniazid
IPT: Isoniazid Preventive Therapy
KII: Key Informant Interview
LTBI: Latent Tuberculosis Infection
MoHCC: Ministry of Health and Child Care
NGO: Non-governmental Organization
NTPSP: National TB Strategic Plan
ORIC: Organizational Readiness for Implementing Change
PLHIV: People living with HIV/AIDS
TB: Tuberculosis
TB – NSP: Tuberculosis national TB strategic plan
TPT: TB Preventive Therapy
WHO: World Health Organization

## Acknowledgements

The authors would like to thank the Ministry of Health and Child Care of Zimbabwe and Clinton Health Access Initiative, Zimbabwe for providing access to personnel involved in the 3HP program under IMPAACT4B. We would also like to thank the respective Bulawayo and Harare City Council Health Service Departments including the facilities for providing access for the collection of data for the study. Thank you to the participants for consenting to partake in the study.

## Consent for Publication

Not applicable

